# Severe Acute Respiratory Syndrome Coronavirus 2 – Reactive Salivary Antibody Detection in South Carolina Emergency Healthcare Workers: September 2019–March 2020

**DOI:** 10.1101/2024.01.31.24301668

**Authors:** Haley C. Meltzer, Jane L. Goodwin, Lauren A. Fowler, Thomas W. Britt, Ronald G. Pirrallo, Jennifer T. Grier

## Abstract

**Background:** On 19 January 2020, the first case of severe acute respiratory syndrome coronavirus 2 (SARS-CoV-2) infection was identified in the US, with the first cases in South Carolina (SC) confirmed on 06 March 2020. Due to initial limited testing capabilities and potential for asymptomatic transmission, it is possible that the virus was present earlier than previously thought while preexisting immunity in at-risk populations was unknown.

**Methods:** Salivary samples from 55 SC emergency department physicians, Emergency Medical Services (EMS) providers, and medical students working as EMTs were collected from September 2019 to March 2020 as part of a separate study and stored frozen. To determine if antibody-based immunity to SARS-CoV-2 was present prior to the first recorded cases, saliva acquired post-shift was analyzed by Enzyme-Linked Immunosorbent Assay with repeat of positive or inconclusive results and follow-up testing of pre-shift samples.

**Results:** Two participants were positive for SARS-CoV-2-reactive salivary IgG, confirmed by repeat and follow-up testing. Positive samples were from medical students working in EMS and were collected in October or November of 2019.

**Conclusions:** The presence of detectable antibodies against SARS-CoV-2 in 2019 suggests that immunity existed in SC, and the US as a whole, prior to the earliest documented cases of COVID-19. Additionally, successful analysis of banked salivary samples demonstrates the feasibility of saliva as a noninvasive tool for surveillance of emerging outbreaks. These findings suggest that emergency healthcare providers represent a high-risk population that should be the focus of infectious disease surveillance.

**Article Summary:** Retrospective SARS-CoV-2 antibody testing of saliva from emergency healthcare workers pre-pandemic identifies two reactive individuals in late 2019. Findings suggest the importance of emergency healthcare workers for infectious disease surveillance and saliva as an effective diagnostic tool.

## Introduction

Since its arrival to the United States (US), severe acute respiratory syndrome coronavirus 2 (SARS-CoV-2), the causative agent of the novel coronavirus disease 2019 (COVID-19) pandemic, has challenged the world’s infectious disease surveillance systems. Its high asymptomatic transmissibility contributed to the rapid international spread that highlighted the extent to which healthcare systems were ill-equipped to manage a novel infectious agent ^1,2^. Efficient, coordinated surveillance of viruses with pandemic potential is an integral aspect of public health and preparedness for outbreaks ^3^.

On 19 January 2020, the first confirmed case of COVID-19 in the US was identified in Snohomish County, Washington ^4^, with the first known case in South Carolina (SC) identified on 06 March 2020 ^5^. However, initial infection reports are likely incomplete due to barriers to accurate viral surveillance. Testing availability in the early months of 2020 was limited due to supply and personnel shortages, as well as an overall lack of knowledge about SARS-CoV-2 ^1,6,7^. Concurrently, individuals with asymptomatic or mild symptomatic cases may not have sought testing, contributing to further infection underreporting ^8–10^. More recent data suggests that the true date of SARS-CoV-2’s arrival to the US was earlier than January 2020. The Retrospective Methodology to Estimate Daily Infections from Deaths algorithm, derived from global seroprevalence and death data, suggests that SARS-CoV-2 may have been in the US in December 2019, and in SC by 30 January 2020 ^6^. Retrospective serological analysis of archived blood samples found the presence of SARS-CoV-2-reactive antibodies in every state tested, suggesting that the virus may have been spreading in the US as early as December 2019 ^8^.

To accurately report the burden of infection in a community, it is important to understand when the pathogen may have been introduced and the prevalence of preexisting immunity. Serological studies have proven effective at revealing the presence of antiviral antibodies that may represent possible infections early during a pandemic ^8,11^. The presence of anti-SARS-CoV-2 antibodies suggests immunity against the pathogen, either occurring from exposure to SARS-CoV-2 or as a result of cross-reactive antibodies arising from an infection with a related pathogen ^12^.

With documented asymptomatic SARS-CoV-2 infections ^13^, testing of only ill patients during the early stages of the pandemic may have contributed to an underestimation of the virus’s prevalence. To obtain a complete picture of SARS-CoV-2 viral burden, it is important to test individuals with and without symptoms. Serological testing of over 7,000 blood specimens detected anti-SARS-CoV-2 antibodies in 1-2% of US donors, with variations in dates and locations of sample collection ^8^. However, this study did not include locations in the Southern US, which became an epicenter for infection burden as the pandemic progressed.

The potential for asymptomatic SARS-CoV-2 transmission also raises concerns with the role that healthcare workers might play in the spread of the virus. Healthcare providers, particularly those with direct patient contact, demonstrated an increased risk of COVID-19 infection compared to the general population ^14^, yet less than 15% of all reported COVID-19 cases in the CDC surveillance system included data on whether the patient was a healthcare worker ^15^. Consequently, COVID-19 exposures, infections, and transmission among healthcare personnel may be largely underestimated.

Selecting the proper surveillance location and population is integral to the effectiveness of early detection and identification of a disease. Emergency healthcare workers (EHCWs) represent one of the first lines of interaction between the community and the healthcare system. Thus, early and focused sampling of Emergency Department (ED) clinicians and Emergency Medical Service (EMS) prehospital providers could contribute to a better understanding of future infectious diseases, particularly those with an asymptomatic, person-to-person transmission. Further, given the interaction between EHCWs and the public with potentially undiagnosed infections, an understanding of existing immunity in this population can provide important insight into their susceptibility.

Early in the pandemic, nasopharyngeal and oropharyngeal swabs were established as the gold standard for SARS-CoV-2 screening. More recently, viral detection in saliva has also demonstrated efficiency and reliability ^16,17^. Benefits of testing salivary samples for infectious diseases and immunity include greater cost-efficiency and lower transmission risk to healthcare workers during sample collection ^18^. Although, accurate and reliable detection of salivary biomarkers may be dependent on collection storage conditions, such as the use of nucleic acid preservation media for detection of viral RNA ^19^. Under the right conditions, saliva is conducive to long-term sample storage, making it an effective tool for retrospective testing.

To determine whether antiviral immunity was present in EHCWs in South Carolina (SC) prior to the first documented cases of COVID-19 and assess the potential of salivary sample testing in immune surveillance, salivary samples collected between September 2019 and March 2020 were retrospectively tested to identify the presence of SARS-CoV-2-reactive IgG.

## Methods

### Setting and Participant Population

Greenville County is located in northwest SC and consists of mixed rural and suburban communities, with the largest city being the City of Greenville. Prisma Health Greenville Memorial Hospital’s ED is the only ACS-verified Adult Level 1 and Pediatric Level 2 Trauma Center in the Upstate, providing emergency care for over 106,000 patients annually.

Study participants included 9 Hospital ED physicians working in an ED ithat includes ACS-verified Adult Level 1 and Pediatric Level 2 Trauma Centers, 7 EMS providers, and 39 medical students working as Emergency Medical Technicians (EMTs) in SC (Table 1).

**Table 1.** Demographic information of participant population.

|  | ED Physicians | EMS Providers | Medical Students |
| --- | --- | --- | --- |
| Number of Participants | 9 | 7 | 39 |
| **SARS-CoV-2 IgG<sup>+</sup>** | 0 | 0 | 2 |
| Gender - % (#) |  |  |  |
| Male | 33.3% (3) | 42.9% (3) | 48.7% (19) |
| Female | 66.6% (6) | 42.9% (3) | 51.3% (20) |
| Other | 0.0% (0) | 14.3% (1) | 0.0% (0) |
| Average Age Range | N.D. | 31-35 | 21-25 |
| **SARS-CoV-2 IgG<sup>+</sup>** | n.a. | n.a. | 25 |
| Shift Duration (hr) | 8 | 12 | 12 |

**Key:** *N.D.* – not determined; *n.a.* – not applicable.

### Human Studies Approval

All salivary samples were collected during two independent studies approved by the Prisma Health Institutional Review Board, in which patient consent allowed for samples to be analyzed for supplementary research. All samples and data were gathered and stored in a de-identified manner.

### Sample Collection and Storage

160 salivary samples were obtained from 55 EHCWs in SC between September 2019 and March 2020. Using the passive drool method, saliva was collected immediately before and after participants’ shifts, which occurred at multiple times of the day and varying days of the week. 1-2 mL of saliva was collected and stored in a 2-4°C refrigeration unit immediately before being moved to a -80°C freezer within 72 hours. Sample testing occurred between January and June 2022 following approximately 2 years of storage.

### Prevention of Contamination

All reactions were prepared in a 1300 Series Class II, Type A2 Biological Safety Cabinet. Prior to use, hood space was UV-treated. Hood surfaces were then sterilized with 10% bleach solution followed by 70% ethanol as were all pipettes and supplies. Only sterile tubes and filtered tips were used. Gloves were changed frequently, and after coming in contact with samples. Personal protective equipment was worn throughout all experiments in accordance with BSL-2 safety protocols.

### ELISA

The presence of IgG against SARS-CoV-2 N protein and the receptor-binding domain (RBD) of the S1 protein was assayed via in vitro, indirect Enzyme-Linked Immunosorbent Assay (ELISA). Frozen salivary samples collected post-shift were thawed at 4°C for 24-48 hours, then vortexed until homogenous and diluted 10-fold. Diluted samples were added to ELISA plates pre-coated with SARS-CoV-2 N/S1 RBD proteins (RayBiotech) and testing was performed according to manufacturer specifications. Bound human IgG antibodies were quantified by optical density (OD) at 450 nm using a microplate reader (Tecan). A simultaneous procedure was performed on human Albumin protein pre-coated ELISA plates, and OD signals were subtracted from N/S1 signals to correct for background interference. Technical duplicates were performed for all samples tested.

For each assay, a calibration curve was calculated and plotted with SARS-CoV-2 N/S1 protein antibody concentration (units/mL) on the x-axis and OD signal on the y-axis. For background-corrected samples, values >0.05 units/mL were used to define a positive result, and values <0.05 units/mL a negative result. For positive or inconclusive results, analysis was repeated on the post-shift sample, and the pre-shift saliva was evaluated across two independent repeats as well. At the time of use, the ELISA kit was labeled for research use only and not FDA-approved for diagnostics.

### Statistical Analysis

Analysis of ELISA data, including calculation of calibration curves, and anti-SARS-CoV-2 N/S1 antibody concentrations were performed according to manufacturer specifications. GraphPad Prism 9.4.1 and Microsoft Excel were used for figure generation and statistical analyses.

## Results

### Participant Characteristics

All participants felt well at the time of sample collection and did not report any respiratory or gastrointestinal symptoms. Participant demographics varied (**Table 1**). Additional demographic information, such as race and ethnicity, was not collected to preserve confidentiality.

Two medical students EMTs were found to produce detectable levels of SARS-CoV-2-reactive IgG, with the presence of antibodies confirmed in two samples obtained from the same individual 12 hours apart. This constitutes a 5.13% positivity rate among the medical student population and an overall participant 3.64% positivity rate. Both participants with SARS-CoV-2-reactive antibodies were female students between the ages of 21-25.

### Detection of SARS-CoV-2-Reactive Salivary Antibodies

All post-shift salivary samples collected from EMS personnel and ED physicians were negative for the presence of anti-SARS-CoV-2 antibodies. A total of 5 post-shift samples from medical students resulted in at least one replicate well testing above the positive threshold and were subjected to follow-up testing. Repeat testing identified two participants positive for SARS-CoV-2-reactive IgG antibodies in both pre-and post-shift salivary samples. Positive Participant 1 (PP1) provided salivary samples in October 2019, and Positive Participant 2 (PP2) provided samples in November 2019 (**Figure 1**).

**Figure 1.**
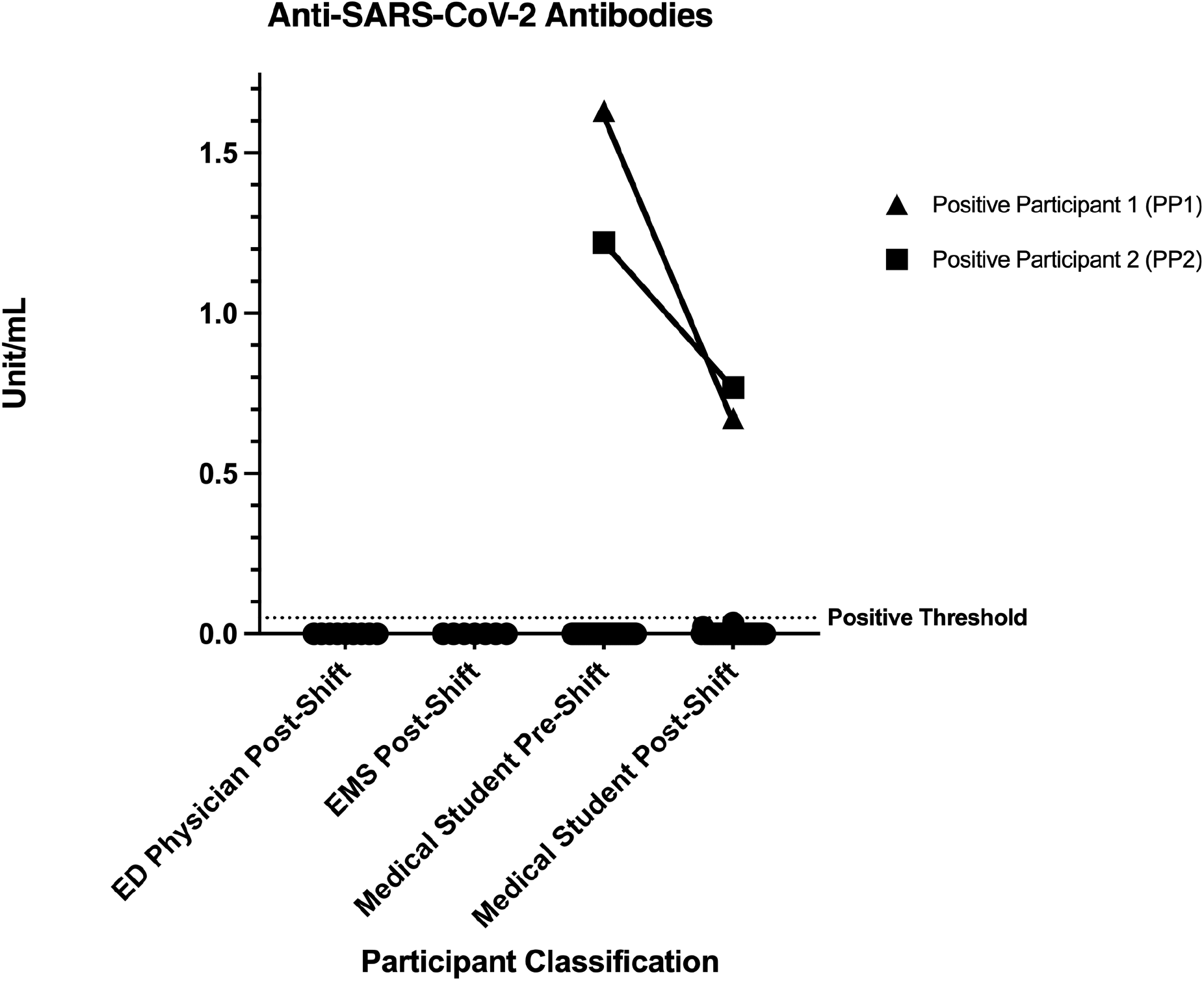
Detection of SARS-CoV-2 S1 RBD- and N-reactive salivary antibodies. Median values are shown. Only samples from PP1 and PP2 were found to have median values above the positive threshold (0.05 unit/mL).

## Discussion

In this secondary analysis of saliva samples collected from EHCWs, we identified antibodies against SARS-CoV-2 in two individuals during the Fall of 2019. Positive results were confirmed in two samples from the same individuals collected 12 hours apart. Repeat testing ruled out potential false positives. This is the first study to detect the presence of SARS-CoV-2 N and S1 RBD reactive salivary IgG prior to locally reported cases of COVID-19.

Serum studies have identified IgG against SARS-CoV-2 S1 in 100% of hospitalized symptomatic patients with COVID-19 by 6 days after hospital admission, and even 36% of patients with asymptomatic infections developed antibodies against the S1 protein ^20^. Furthermore, patients with asymptomatic or mild SARS-CoV-2 infections produced significantly higher levels of serum IgG against both the S1 and N viral proteins compared to exposed healthy controls ^21^. Together, these studies suggest that the presence of IgG against SARS-CoV-2 observed in the current study indicates a potential for a prior coronavirus infection, rather than simply exposure to infected patients during healthcare work. Based on the timing of positive sample collection in October and November 2019, SARS-CoV-2 IgG-positive individuals in this study may have experienced SARS-CoV-2 infection prior to the earliest documented cases in the US. Participants could not be retrospectively queried about possible exposure history; thus, it is not possible to determine if an infection occurred as a result of travel or community acquisition. While positive participants were asymptomatic at the time of sample collection, the detection of SARS-CoV-2-reactive antibodies in these salivary samples supports the idea that EHCWs and students who provide emergency care could be an optimal population for surveillance of emerging respiratory infections.

### SARS-CoV-2-Reactive Salivary IgG

The detection of anti-SARS-CoV-2 antibodies in saliva has been presented as an alternative approach to serum testing for surveillance of antiviral immunity in a population ^22–26^. The use of salivary samples for detection of antiviral antibodies has been successfully implemented for a variety of viruses, including Hepatitis A and HIV, and provided the groundwork for salivary SARS-CoV-2 testing ^27–30^. Studies focused on antibody responses during and after infection have validated the correlation between serum and salivary antibody levels ^22,23,31^. Advantages of using saliva samples include the ease of sample collection and the ability to store whole samples for future testing, whereas serum samples require isolation from whole blood prior to long-term storage ^32^. Additionally, saliva can be collected by an individual in a noninvasive manner, reducing exposure risks to healthcare workers relative to collection of serum samples, which require close contact with a potentially infectious patient.

Salivary antibodies against either S1 RBD or N protein were rarely detected in individuals with a COVID-negative history, despite high rates of antibody reactivity with antigens from common coronaviruses in the same population ^23^. IgG reactive against SARS-CoV-2 S1 and N have been identified in the serum of uninfected patients with samples collected as early as 2011, suggesting that cross-reactive antibodies may be generated during infection with other human coronaviruses ^33–35^. A greater percentage of young patients (6-16yrs) were found to carry serum IgG antibodies against SARS-CoV-2 compared to young adults (17-25yrs), which correlates with the increased likelihood of human coronavirus (HCoV) infection and seroconversion during adolescence ^33^. The 5.13% rate of SARS-CoV-2-reactive antibodies in this study fell below the previously reported serum detection range of 5.72-43.75% for patients over the age of 17 ^33^. However, this is likely a result of limited participant numbers and the use of alternate samples for antibody detection. Interestingly, SARS-CoV-2-negative patients with higher levels of preexisting IgG against HCoV N protein had lower levels of SARS-CoV-2 N-reactive IgG, indicating a potential protective association between prior HCoV infection and risk of SARS-CoV-2 infection ^34^. Furthermore, SARS-CoV-2 S-reactive antibodies from pre-pandemic serum samples proved capable of neutralizing infection by SARS-CoV-2 S pseudotypes or native SARS-CoV-2 infection ^33^. This supports the conclusion that participants in this study who tested positive for the presence of SARS-CoV-2-reactive IgG likely possessed some immunity against infection.

This study assessed the presence of IgG antibodies that bind either the S1 RBD or N protein of SARS-CoV-2 in the saliva of EHCWs who may have been at risk of exposure in the earliest stages of the pandemic. Antibodies against these viral components are routinely detectable in saliva following SARS-CoV-2 infection ^24,26^. While IgA and IgM antibody levels in saliva decay shortly after infection, IgG antibodies against SARS-CoV-2 persist and remain detectable up to 9 months post-infection, demonstrating a reliable method of detecting potential prior infections ^23,26^. In addition, salivary antibodies may also serve as an indication of immunity against SARS-CoV-2. Salivary antibodies can directly contribute to immune protection and given the strong correlation between salivary and serum antibody levels, they can signify the presence of neutralizing antibodies in the serum ^25^. Detection of SARS-CoV-2-reactive salivary antibodies in two participants in this study demonstrates that a small percentage of EHCWs may have had preexisting immunity to SARS-CoV-2 prior to the first documented local cases. Understanding the prevalence of immunity in a population, regardless of whether that immunity arose from prior infection or cross-reactive antibodies, is an important step in evaluating susceptibility to infection and potential for contribution to viral transmission.

A limitation of this study is that we cannot confirm the presence of true positives for SARS-CoV-2 via a positive molecular test for detection of viral components, such as viral RNA via PCR test ^8,36^. The samples with confirmed positive ELISA results for the presence of anti-SARS-CoV-2 antibodies cannot be definitively considered positive for SARS-CoV-2 infection without confirmation by molecular diagnostic testing ^8,36^. Unfortunately, molecular analysis for SARS-CoV-2 RNA in these samples has not proven successful, despite multiple approaches and attempts. One potential reason why RNA testing may have failed is due to sample collection and storage conditions. PCR detection of viral RNA can be inhibited by the presence of blood, mucus, or food particles in saliva ^37^; thus, it is advisable that participants avoid eating or drinking in the 30 minutes prior to sample collection. Due to clinical care duties, it is unknown if this waiting period occurred. Additionally, as the samples were not collected with the intention of detecting RNA, they were not stored in a nucleic acid preservation storage media. Additionally, sources of samples were anonymous, and further history or exposure risks could not be assessed.

## Conclusions

The exact date of SARS-CoV-2’s arrival to the US may never be known; therefore, understanding the level of immunity in the population is essential to inform surveillance measures and public health decisions. Considerations in infectious disease surveillance highlight the significance of EHCWs as a high-risk population for exposure, infection, and potential transmission. Our data offers a unique glimpse into SARS-CoV-2 immunity in EHCWs during the time prior to reported cases and suggests that EHCWs themselves could be a focus of disease surveillance and detection of immunity. Furthermore, the ability to detect pathogen-reactive antibodies in saliva, and challenges with successfully completing molecular analysis, emphasizes the importance of proper collection and storage for viral detection. This work establishes the potential for screening programs in EHCWs to identify protective immunity and determine disease susceptibility in this high-risk population.

## Data Availability

All data produced in the present study are available upon reasonable request to the authors.

## Acknowledgements

We would like to acknowledge the following for their contributions to the collection of samples used in this study: Anjali Amalean, Kyleigh Connolly, Haley Fulton, Haritha Pavuluri, W. Michael Schmidt (University of South Carolina School of Medicine Greenville); Kirby Allen, Alyssa Banks, Nicole Banks, Caroline Barrows, Alexxa Bessey, Madison Faulkner, Sophie Finnell, Nicole Fuller, Madison Gassmann, Courtney Gouge, Cassidy Graham, Luke Hall, Nancy Hillman, Rebecca Kerr, Zachary Klinefelter, Brady Kluge, Nabihah Kumte, Riley McCallus, Melissa McIntee, Zoey Morten, Cavan Peters, Kaustubha Reddy, Laura Scherck, Claudia Stohr, Bailey Taylor, Delaney Wallace, Brianna Weiss, Ann Louise Williamson, and Chloe Wilson (Clemson University).

## Author Contributions

The study was conceived by LAF and JTG with experimental design contributions from TWB and RGP. Data acquisition was performed by HCM and JLG, and HCM conducted data analsyis. HCM and JTG drafted the manuscript with revisions provided by HCM, LAF, TWB, RGP and JTG.

